# Single-cell analysis of follicular fluid reveals dysregulation of ovulatory immune function in PCOS patients undergoing ovarian stimulation

**DOI:** 10.1101/2025.08.11.25333027

**Authors:** Andrea K. Wegrzynowicz, Soma Banerjee, Emily Grimes, Riley Huddleston, Fernanda Leyva Jaimes, Laura G. Cooney, Aleksandar K. Stanic

## Abstract

Polycystic ovary syndrome is the most common endocrine condition in women and anovulatory cause of female infertility. While a pro-inflammatory cytokines and leukocyte bias in systemic circulation is well-documented in PCOS, it is not known how this inflammation extends to or affects the ovary. Additionally, the relationship between ovulation and inflammation in PCOS is not well-defined. We hypothesize that the ovarian follicular immune environment in PCOS is uniquely dysregulated, and that resolving anovulation through ovulation induction is not sufficient to alleviate this dysregulation. Using single-cell RNA and surface protein analysis of peripheral blood and follicular fluid from patients undergoing *in vitro* fertilization, we discovered that both control and PCOS follicles were immunologically distinct from circulation. At a systemic level, we find that ovulation induction in PCOS does not alleviate systemic inflammation. In contrast, while healthy control ovaries experienced acute immune-directed ovulatory signaling, PCOS ovarian follicles were deficient in key pro-ovulatory cell to cell communication, and displayed instead a chronic low-grade inflammatory state with fibrotic features. Taken together, a picture emerges where acute ovulation demonstrates a well-ordered series of follicle-specific immune information flows, which are disrupted and replaced by low grade chronic inflammation in the PCOS follicle.

## INTRODUCTION

Polycystic ovary syndrome (PCOS) is the most common endocrine disorder, affecting millions of reproductive-aged individuals with ovaries and making it one of the most common causes of female factor infertility.^1^ PCOS is defined by the Rotterdam Diagnostic Criteria as the presence of 2 out of 3 of the following - 1) Clinical or biochemical hyperandrogenism, 2) Oligomenorrhea, and 3) Polycystic ovaries on ultrasound.^1^ Visceral obesity, hyperinsulinemia, and insulin resistance are also common in PCOS, further influencing reproductive and metabolic health.

PCOS is associated with low-grade systemic inflammation, including elevation of inflammation-related cytokines (TNF-α, IL-6, MCP-1 (CCL2), MIF, IL-18, and IL-1b)^2^ and higher M1/M2 macrophage and Th17/Th2 ratios.^2–8^ While the inflammation associated with PCOS is well-characterized in circulation, the specific inflammation experienced by the ovarian follicle as well as the cellular sources of inflammatory molecules is not defined. Follicular fluid, extracted at the time of transvaginal oocyte retrieval during in vitro fertilization (IVF) has shown higher levels of pro-inflammatory markers (TNF-α, CRP, IL-6, IL-2 and IFNγ) in individuals with PCOS compared to controls,^9–11^ but the relationship between systemic inflammation, ovarian inflammation, and ovulation is not clear.

This is significant as normal ovulation is a controlled inflammatory process, with release of pro-inflammatory cytokines and chemokines and recruitment of cells including macrophages, monocytes, and T cells to the follicle immediately before ovulation.^12,13^ Given potential overlap between this acute inflammatory environment in the peri-ovulatory ovary and the chronic inflammation observed in PCOS, we sought to characterize the immune environment in the ovarian follicle at a single-cell level in both control and PCOS patients. We hypothesized that PCOS follicles would display unique immune dysregulation, differing from both control follicles and from the systemic inflammation observed in PCOS.

To test this hypothesis, we collected blood and follicular fluid at the time of transvaginal oocyte retrieval in patients undergoing *in vitro* fertilization at a university-associated fertility clinic. We used CITEseq (Cellular Indexing of Transcriptomes and Epitopes) to sequence and characterize mononuclear cells in both the ovarian follicle and peripheral blood.^14^ This led to identification of important inflammatory pathways in normal ovulation and disruption of these pathways in the PCOS follicle even as systemic inflammation persisted. Our data suggest that ovarian inflammation and immune dysregulation are not due to anovulation alone and that the PCOS follicle has a distinct immune profile.

## RESULTS

### Unique immune environment of the ovulatory ovarian follicle

8 patients were included in our study: two each in the non-obese PCOS, obese PCOS, non-obese control, and obese control groups (Table 1). Patients were age- and BMI-matched, with patient identifiers increasing with BMI (i.e. PCOS A and Control A are lowest BMI and PCOS/Control D are the highest). Following isolation and sequencing,^14^ 68601 cells passed quality control metrics (Figure 1A, Figure S1). Cells were first clustered based on canonical immune marker genes into relatively broad categories (Figure 1B, C), with T cells being the most populous cell type (Figure 1D). T cells, NK cells, B cells, Monocyte lineage cells, and follicular cells were then further subclustered for major known immune cell subsets based on feature barcoded antibody levels and canonical gene expression for all cell types (Figure 1E-G, Figure S2)^15–18^.

**Table 1.** Study groups and inclusion/exclusion criteria.

|  | Group 1 | Group 2 | Group 3 | Group 4 |
| --- | --- | --- | --- | --- |
| Patient Population | Healthy Control Patients (infertility due to non-PCOS associated causes) |  | PCOS Patients |  |
| BMI | BMI <30 | BMI >30 | BMI <30 | BMI >30 |
| Inclusion Criteria | Regular menstrual cycles, normal ovarian reserve, age 18-44 |  | Meets Rotterdam Criteria |  |
| Exclusion Criteria | Current pregnancy, endometriosis, diabetes, immunomodulating medications, autoimmune conditions, recurrent pregnancy loss, active malignancy, in-situ hydrosalpinx without ligation |  |  |  |

**Figure 1.**
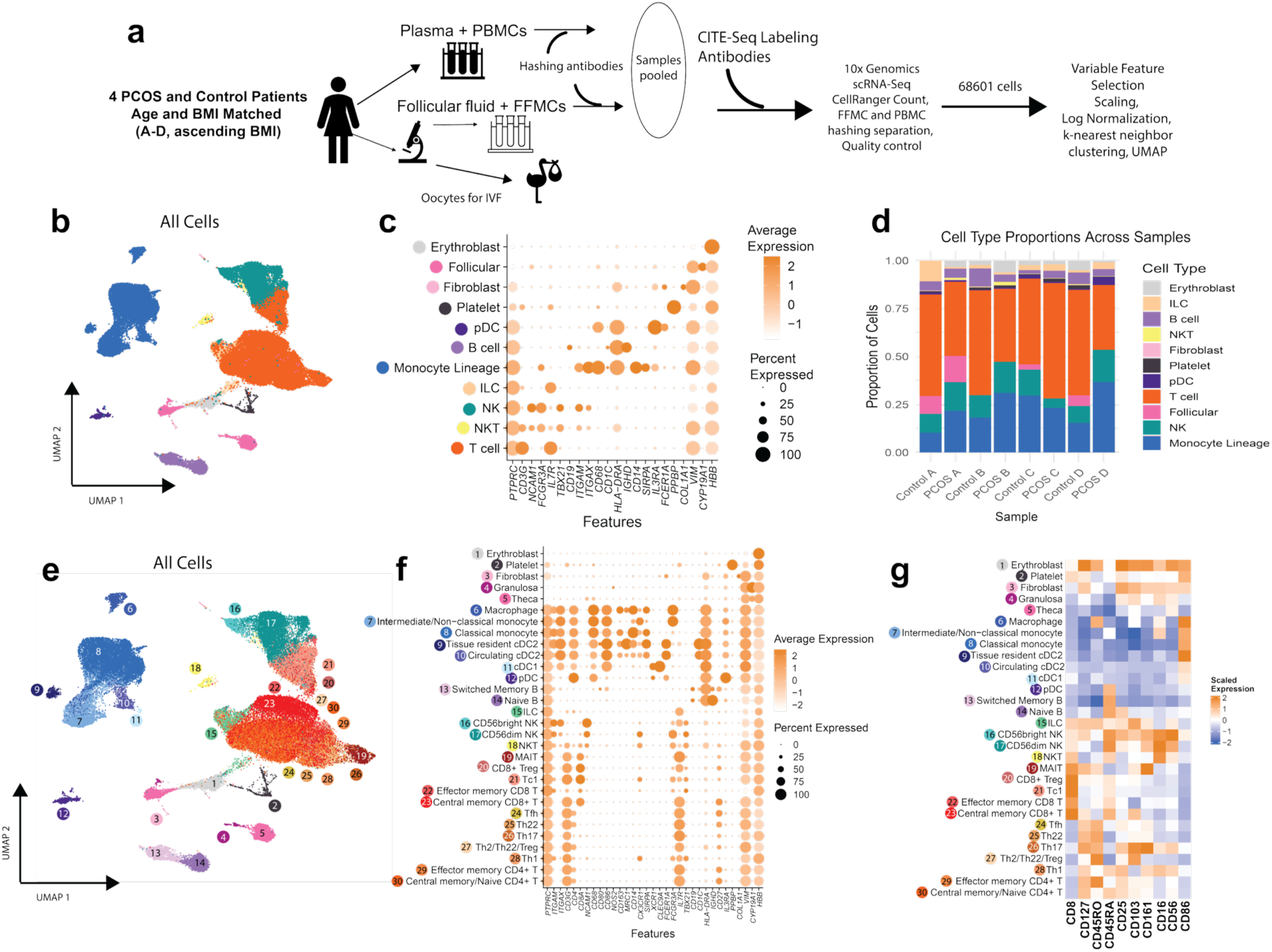
Identification of peripheral blood mononuclear cells and follicular fluid mononuclear cells by transcriptomic and proteomic markers. A) Blood and follicular fluid were collected from 4 PCOS and 4 Control patients, and mononuclear cells isolated and sequenced via CITEseq. Cells passing quality control were passed through our previously used analysis pipeline and unsupervised clustering was performed via UMAP at resolution 0.8. B) UMAP clusters were annotated as broad cell types, based on C) expression of canonical immune-subset defining genes. D) Overall cell type proportions by patient (blood and follicular fluid combined). E) Following subclustering, cell identities were annotated with greater precision based on F) gene expression and G) proteomic analysis.

We combined all cells (PBMCs and FFMCs, in both PCOS and control patients) for clustering and annotation. Following annotation, we compared these conditions (Figure 2A) in order to determine if follicular fluid is a distinct immune compartment and whether PCOS impacts its cellular composition. We found that major cellular categories were similar (Figure 2B), indicating that the ovarian follicle is not a dramatically distinct, immune privileged environment. That said, ovarian follicle is a uniquely regulated immune environment, as demonstrated by a set of specific cellular differences compared with matched systemic circulation. We discovered that classical monocytes were lower, while macrophages, Th22 subset, and tissue-resident cDC2s were all enriched compared with matched PBMCs of the same individuals (Figure 2C, Table S4). We next tested the hypothesis that immune cellular composition is dysregulated by PCOS. Our data however suggest that PCOS and controls are closely matched, with the singular exception of tissue-resident cDC2s which were highly enriched in PCOS patients (4.5% vs. 1.3%, p = 0.026) (Figure 2D, Table S4). While our experimental design was not geared towards specific isolation of granulosa and theca cells (see Methods), a small proportion overall was identified in samples, and were by definition only present in the follicular fluid. Notably, there were no statistically significant differences between PCOS and controls of proportions of immune subsets in peripheral blood between PCOS and control patients (Figure S3). Consequently, the immune environment of the follicle is distinct compared with circulation, with specific immune cell subsets recruited to or resident in this environment, and with PCOS only directly impacting the proportion of follicular cDC2s. These data suggest that systemic (circulation) inflammation seen in PCOS is not driven by enrichment of lymphocyte subsets, but instead is driven by programming, a hypothesis we set out to test next.

**Figure 2.**
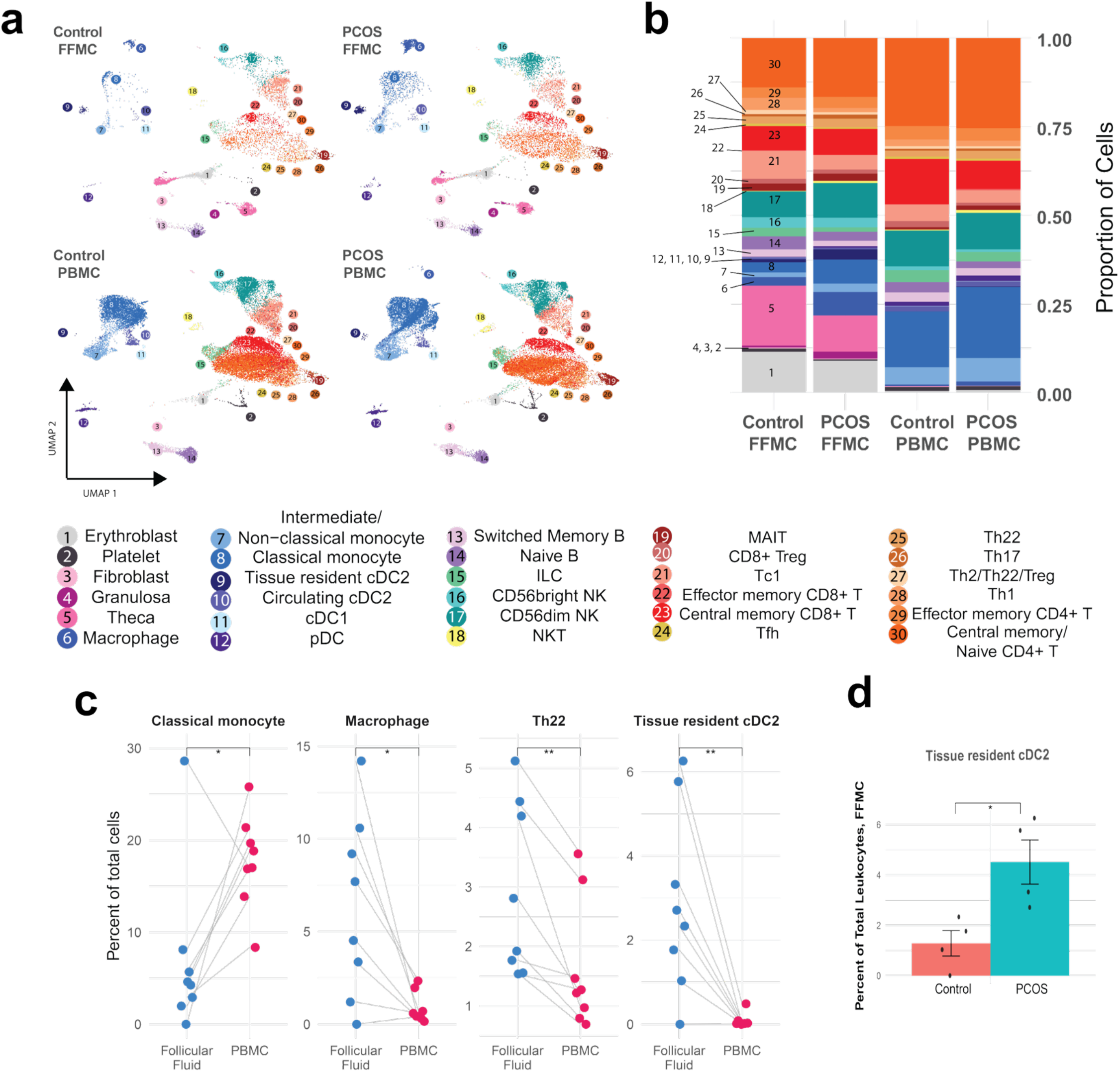
Follicular fluid mononuclear cell composition varies slightly from peripheral blood. A) Cell identities in follicular fluid (top) and peripheral blood (bottom), compared in control (left) and PCOS (right). B) Cell identity proportions in all four conditions. C) Classical monocytes are of a greater proportion in PBMCs, while macrophages, Th22, and tissue-resident cDC2 cells are of a greater proportion in FFMCs. D) Tissue-resident cDC2s are elevated in PCOS patients compared to controls. Statistical analyses: Student’s t test, * p < 0.05, ** p < 0.01.

### Ovulation induction for IVF does not alleviate systemic inflammation in PCOS patients

While systemic inflammation is well-documented in PCOS, the cellular origins of these inflammatory mediators and their status in the setting of ovarian stimulation is less defined. If anovulation alone is the cause of inflammation in PCOS, we hypothesized that this inflammation should be absent in our PCOS patients undergoing successful ovulation induction for IVF. On the contrary, our results reveal that peripheral immune complement in PCOS displays a unique immune transcriptome compared to controls (Figure 3A), notable for overexpression of major histocompatibility type II transcripts (fold change > 4, FDR = 0, antigen presentation, upregulated in inflammation), with a number of downregulated genes broadly associated with cell metabolism and proliferation (e.g, CPA5, SLC35F1, DHCR24, negative fold change > 4, FDR ≈ 0). To better determine the functional groupings of upregulated transcripts in PCOS, we analyzed the entire gene dataset for enrichment of Gene Ontology (GO) terms. GO analysis was very revealing, indicating an overall inflammatory profile, including upregulation of genes involved in the inflammatory response, cytokine production, TNF production, and leukocyte activation (Figure 3B). Notably, genes were not driven by patient subgroups as the inflammatory response was overexpressed in all PCOS patients compared to controls (Figure 3C). Finally, we looked for the cellular sources of these PCOS inflammatory signals, and found that monocyte-derived cells (especially macrophages) were largely responsible for expression of TNF, IL18, IL-1B, CCL2, CCL3, and IL10. Natural killer T cells (NKTs, CD3+CD56+) and Th17 cells, both powerful effector subsets, displayed pro-inflammatory transcriptional programming (Figure 3D). Overall, these results confirm that PCOS systemic inflammation is not corrected by successful ovulation induction, indicating that this is either an independent process, or requires a long-term sustained ovulatory physiology to normalize.

**Figure 3.**
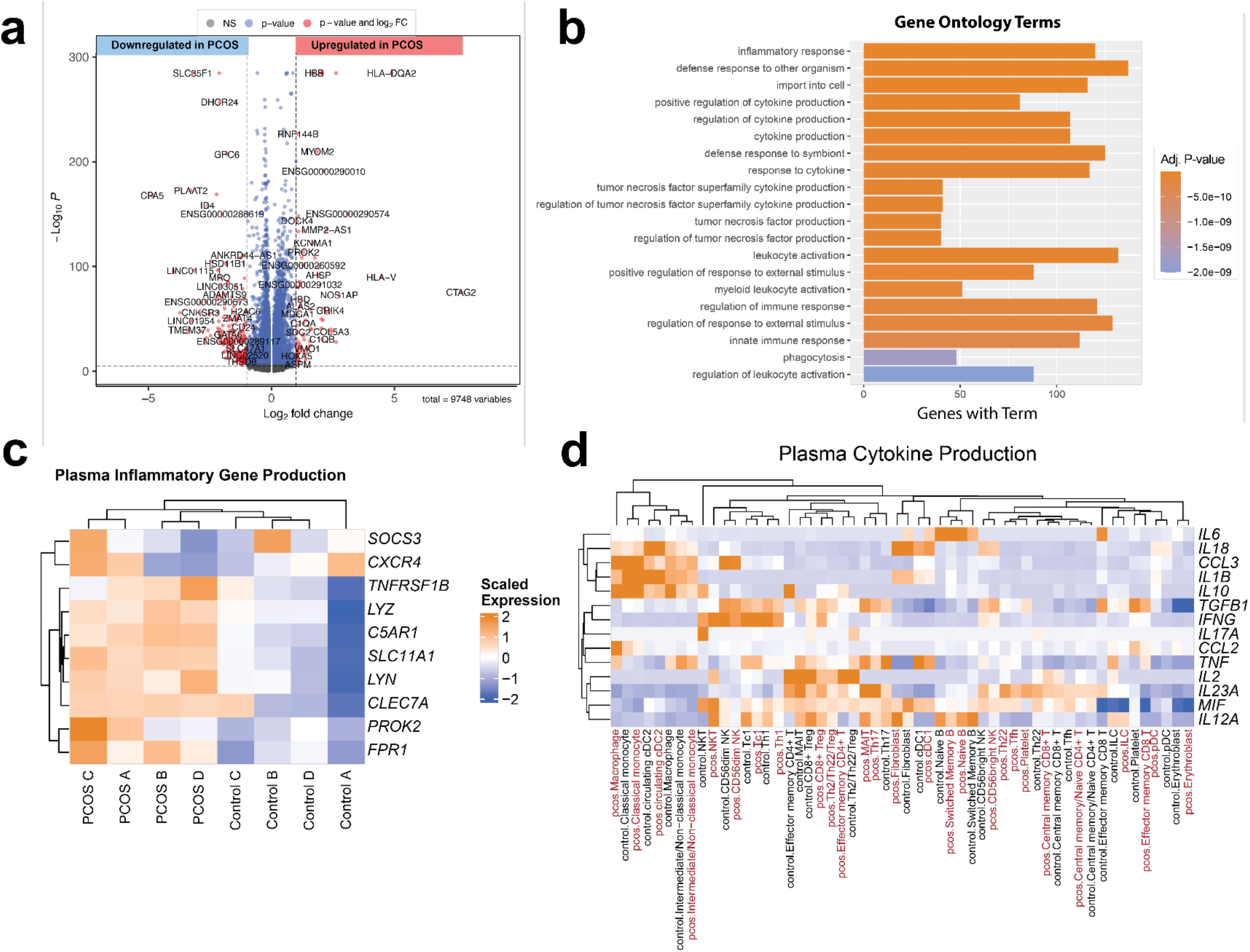
Pro-inflammatory pathways and cytokines are upregulated in peripheral blood of PCOS patients undergoing IVF. A) Differential gene expression analysis of genes upregulated (positive log2FC) and downregulated (negative log2FC) in PCOS PBMCs compared to controls. B) Top 20 Gene Ontology terms of genes upregulated (log2FC > 0.25, FDR < 0.05) in PCOS PBMCs compared to controls. C) Mean expression of top 10 genes annotated as “inflammatory response” in all patients. D) Mean expression of cytokines known to be elevated in peripheral blood of PCOS patients, by cell identity and PCOS status.

### Orchestrated immune activity characteristic of ovulatory follicle is attributable to monocytes, macrophages, and conventional dendritic cells

To understand how immune dysregulation in PCOS presents in the ovary specifically, it is important to determine the immune processes of *normal* ovulation. Ovulation is a choreographed acute inflammatory process, with the LH surge resulting in infiltration of leukocytes, particularly macrophages and monocytes, as well as production of chemokines and cytokines including CCL2, CCL20, TNF, PGF, and IL-1B.^19–26^ To determine immunocellular drivers of these events in healthy controls we first determined the overall follicular leukocyte transcriptome compared with matched peripheral blood and observed the expected acute inflammatory signature. Notable transcriptionally active genes compared to circulation were CCL2, TNF, IFNG, and CXCL2 (Figure 4A). KEGG pathways related to inflammation were highly upregulated, including cytokine-cytokine receptor interaction, antigen processing, and the TNF signaling pathway (Figure 4B).

**Figure 4.**
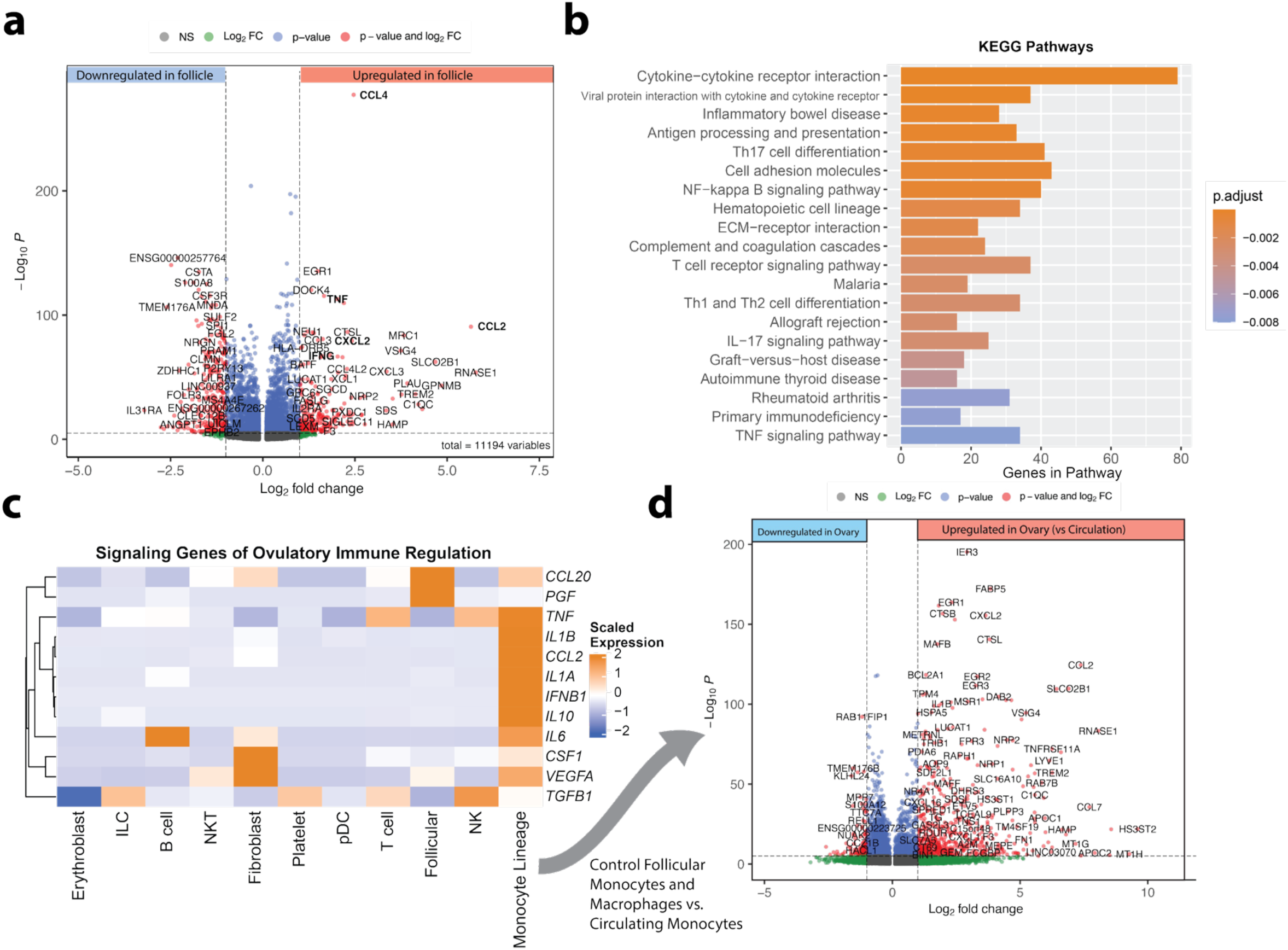
The peri-ovulatory follicle is an active immune environment due largely to activity of monocytes, macrophages, and conventional dendritic cells. A) Differential expression analysis of genes upregulated (positive log2FC) and downregulated (negative log2FC) in control FFMCs compared to PBMCs. B) Top 20 KEGG Pathways of genes upregulated (log2FC > 0.25, FDR < 0.05) in control FFMCs compared to PBMCs. C) Mean expression of signaling genes important for physiological ovulation. D). Differential expression analysis of genes upregulated (positive log2FC) and downregulated (negative log2FC) in control follicular monocytes and macrophages compared to the same cell types in circulation.

Focusing on the cellular sources of cytokines and chemokines known to be important for ovulation, we found that vast majority were expressed by monocytes and monocyte-derived cells, and a few by non-immune follicular (granulosa, theca, etc.) cells (Figure 4C). We specifically examined the differential transcriptional programming of follicular monocytes and macrophages (compared to circulating, Figure 4D) and found that it captured most of the immune activity related to ovulation. We noted a few other immune populations with minor differences in follicular gene expression compared to circulation, including Tc1 and CD56dim NK cells (Figure S4). Taken together, our high-resolution analysis confirms and extends the current understanding of monocyte/macrophage lineage as primary orchestrators of normal immune ovulatory activity.

### Normal immune activity is dysregulated in follicular fluid of PCOS patients

Next, we asked if correcting the ovulatory defect in PCOS by external FSH, LH and trigger also restores the normal acute ovulatory immune reaction. To systematically dissect this question, we first analyzed differential gene expression between PCOS circulatory and follicular lymphocytes, which revealed a disorganized immune activation (Figure S5A), indicating a failure to restore the normal response (Figure S5A and S5B) by successful ovulation induction in PCOS. KEGG pathways related to autoimmune disease were also upregulated in the PCOS ovarian follicles, consistent with chronic low-grade inflammation (Figure S5B). Noting the immune dysregulation in PCOS follicular immune cells compared to periphery, we next determined if the pivotal cytokine mediators of PCOS-related inflammation seen in the periphery are also matched in follicular fluid. To our surprise, we found that these were in fact most highly expressed in *control* follicles, and the PCOS follicles displayed the lowest expression (Figure 5A) suggesting a displacement of the inflammatory cytokine production from the follicle to periphery in PCOS. While many of these genes are also important to ovulation, we did not expect to find that they were downregulated in PCOS, despite upregulation of inflammation-related gene ontology terms more broadly.

**Figure 5.**
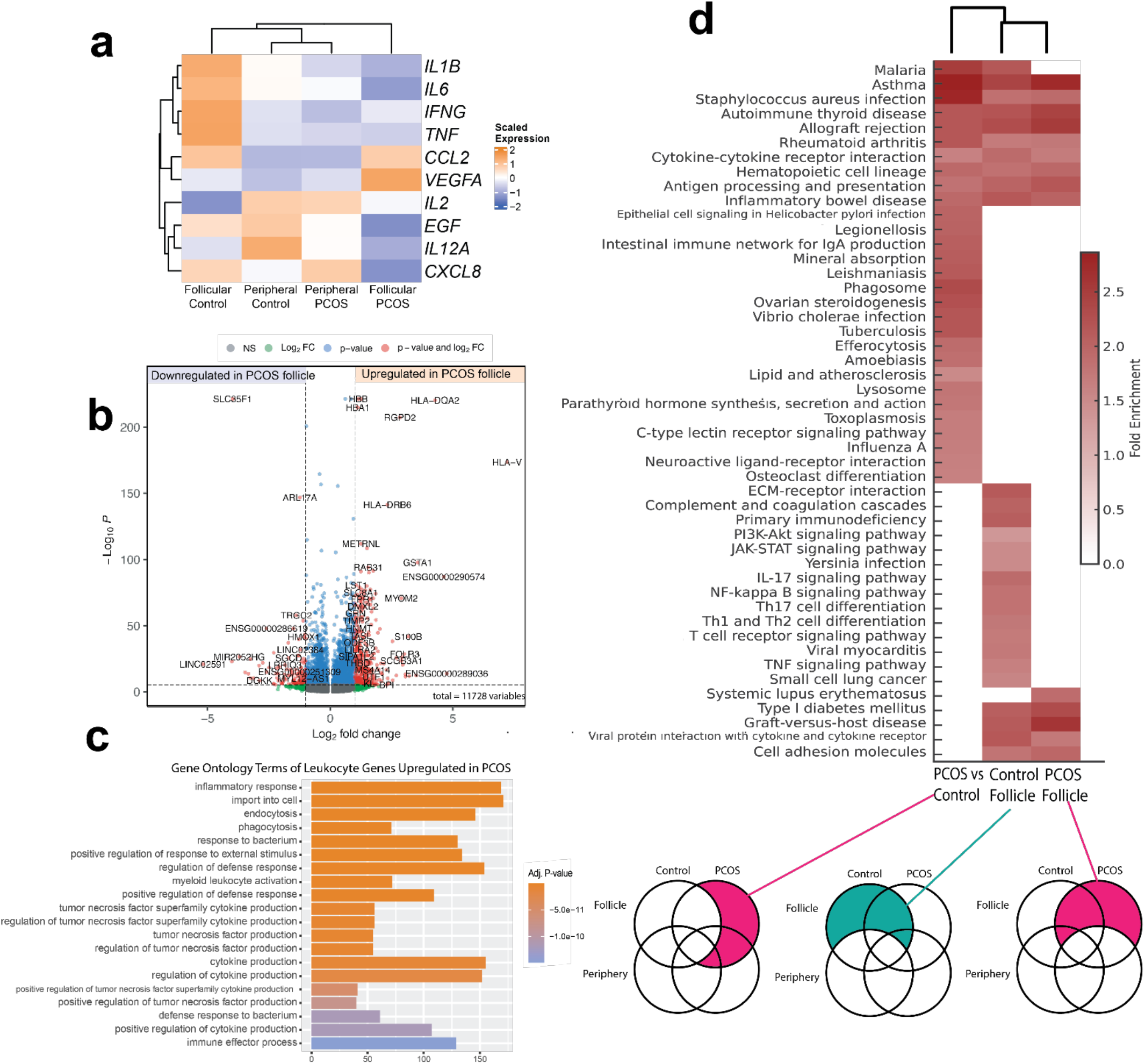
Generalized inflammation is upregulated in PCOS follicles compared to controls. A) Mean expression of cytokines typically elevated in PCOS periphery. B) Differential expression analysis of genes upregulated (positive log2FC) and downregulated (negative log2FC) in PCOS FFMCs compared to controls. C) Top 20 Gene Ontology terms of genes upregulated (log2FC > 0.25, FDR < 0.05) in PCOS FFMCs compared to controls. D). Fold enrichment of top 15 KEGG pathways found in the gene sets that are upregulated in PCOS follicles compared to control, control follicles compared to periphery, and PCOS follicles compared to periphery.

These data raised the question of how different is immune transcriptional programming in PCOS compared with healthy follicle (differential gene expression, Figure 5B) and subjected them to GO term matching (Figure 5C). Enriched gene sets in PCOS related to inflammatory response, namely regulation of responses to external stimulus, immune activation, and cytokine production (Figure 5C).

To better interpret these conflicting results and expression of inflammation-associated genes by both the PCOS and control cells, we examined the most highly expressed (log2FC >1) and significantly upregulated (FDR < 0.05) genes shared by and *unique to* the follicles of PCOS and control patients, compared to their respective periphery and to each other (see schematic below Figure 5D). We then carried out KEGG pathway analysis of the lists of overlapping and unique upregulated genes, and found that, while many immune-related pathways were enriched in all the follicular leukocytes, there were some that were unique to PCOS and others unique to controls (Figure 5D). In particular, pathways related to infectious disease and lipid regulation were upregulated in PCOS follicles compared to controls. Control follicles were uniquely enriched for signaling pathways including JAK-STAT, IL-17, NF-κB, and TNF, whereas PCOS follicles were uniquely enriched for pathways related to autoimmune diseases.

Upon examining the genes in the “inflammatory response” KEGG pathway that were highly expressed in both PCOS and control follicular cells compared to their peripheral counterparts, we found that control macrophages displayed higher expression of these genes overall (Figure S5C). Additional notable findings were selective genes expressed by theca and fibroblasts, CD56bright NK cells, and tissue resident cDC2s. PCOS NKTs and CD8+ Tregs in particular displayed higher expression of CCL4, IFNG, and CCL18 than their control counterparts. Conversely, control classical monocytes displayed a more inflammatory expression profile than the PCOS classical monocytes. Taken together, this suggests that while there is overall immune activity required for ovulation, this immune activity remains tightly regulated in the control follicles, whereas in PCOS follicles there is a more generalized and dysregulated inflammation consistent with the chronic systemic inflammation observed in PCOS.

### Ovulatory signaling pathways are disrupted in macrophages and monocytes of PCOS follicles

As our results suggested that several key immune signaling pathways were absent or downregulated in the PCOS follicles, we next asked if cell-cell information flow was disrupted in PCOS. We employed CellChat to evaluate how cell-cell receptor to ligand interactions compared between the PCOS and control datasets.^27^ We found that the TNF pathway had the highest information flow in control compared to PCOS, consistent with its identification as an upregulated pathway in controls as well as its role as a key pathway in ovulation (Figure 6A).^23–25^ VEGF, IL-1, and CCL signaling were similarly upregulated in controls. Most importantly, IL-10 signaling was also upregulated in controls, indicating that anti-inflammatory balance is also necessary for normal ovulation. When these cell-cell communication pathways were examined in detail (Figure 6B), TNF and VEGF signaling were entirely absent in the PCOS dataset, and in CCL, IL-1, and IL-10 pathways, macrophage and monocyte signaling in particular were disrupted in PCOS compared to controls.

**Figure 6.**
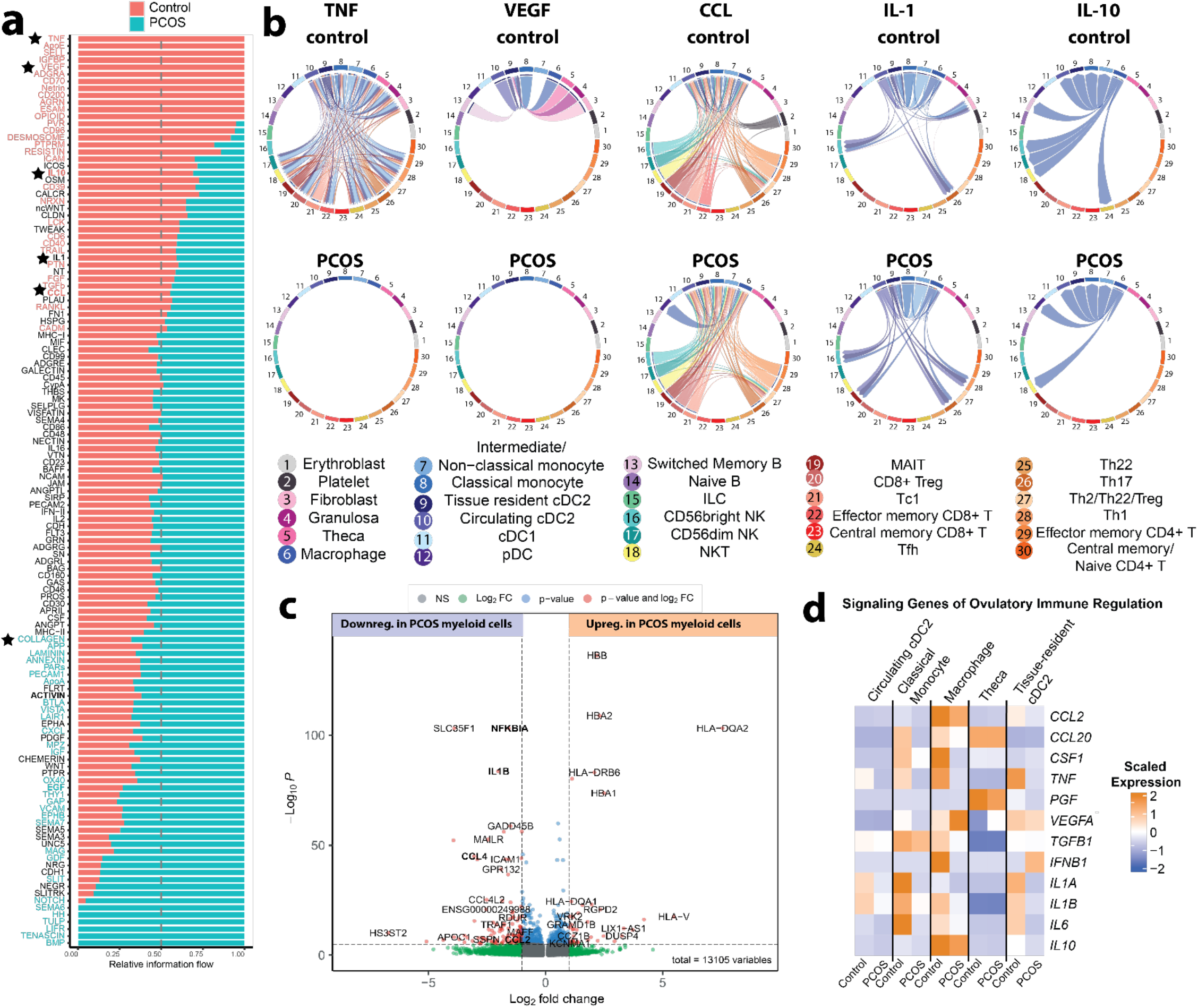
TNF, IL1, IL10 and CCL signaling pathways are dysregulated in PCOS follicles. A) Relative information flow of all signaling pathways detected in PCOS and control datasets by CellChat. Stars indicate pathways discussed further. B) Signaling pathways with sending and receiving cell types specified. TNF and VEGF pathways were not detected in PCOS samples. C) Differential expression analysis of genes upregulated (positive log2FC) and downregulated (negative log2FC) in PCOS monocytes and macrophages compared to controls. D) Mean expression of ovulation-related immune signaling genes by select myeloid subsets.

To further confirm the role of macrophage and monocyte signaling in PCOS-related immune dysregulation, we carried out differential gene expression of PCOS monocytes and macrophages compared to control monocytes and macrophages, and found that NFκB, IL1B, and CCL4 were significantly downregulated, consistent with the CellChat and previous KEGG pathway results (Figure 6C). Finally, signaling genes known to be important in ovulation (see also Figure 4C) were specifically downregulated in PCOS classical monocytes, macrophages, and tissue-resident cDC2s (Figure 6D). Analysis of other cell types confirmed that differences in gene expression between PCOS and control follicles were primarily confined to monocytes and macrophages (Figure S6).

## DISCUSSION

While chronic inflammation in PCOS in the systemic compartment is well understood, the tissue-specific ovarian inflammation and its relationship to normal ovulation is less clear.^2^ Here, we hypothesized that PCOS follicles have unique immunological dysregulation, differing from both control follicles and from the systemic inflammation observed in PCOS. Due to the nature of our study population, collecting follicular fluid from IVF patients, we were also inherently testing if this systemic inflammation was present *during stimulated ovulation*, and therefore if PCOS-related inflammation was independent of anovulation.

We annotated 30 cell identities within follicular fluid, contributing to our understanding of the peri- ovulatory immune environment. While our study was not powered to identify small differences in abundance of different immune cell populations, we did identify that macrophages and Th22 cells were more abundant in follicular fluid, and classical monocytes were less abundant (as expected, since monocytes primarily reside in circulation). Notably, we identified a tissue-resident cDC2 population that was both more prevalent in PCOS and transcriptionally distinct from their circulating counterparts. Despite cDC2s being more abundant in PCOS, their ovulatory gene expression modules were turned off, indicating disrupted function.

In control follicles, multiple inflammatory signaling pathways were robustly upregulated, consistent with the requirements of ovulation. Signaling pathways such as TNF, IL-1, and NF-kB were strongly activated in the control peri-ovulatory leukocytes (Figures 4 and 6), consistent with the surge of cytokines and leukocyte recruitment triggered by the LH surge.^12^ This pro-inflammatory cascade essential for follicular rupture and luteinization is acute and must be tightly regulated, and we also observed upregulation of the counter-regulatory IL-10 signaling pathway in control follicles. This suggests that the control stimulated ovulation is tightly balanced at the immune level. Monocytes, macrophages, and cDC2s emerged as the central orchestrators of this regulation, consistent with prior evidence of their involvement in ovulation and subsequent tissue remodeling.^12,28–32^ This is further supported by increased expression of CCL2 (MCP-1) in ovarian follicular cells, recruiting monocytes to the area.

In contrast, PCOS follicles demonstrated a different profile more consistent with chronic low-grade inflammation, despite stimulated ovulation. We found enrichment of pathways related to extracellular matrix organization and fibrosis (i.e. increased collagen and laminin signaling; Figure 5D). This fibrotic microenvironment may impede normal leukocyte migration, as typically collagen is broken down by matrix metalloproteinases during the ovulatory process.^12,33^ and indeed ovarian collagen deposition is a known feature of PCOS.^34–36^ Our results are consistent with prior studies that have found upregulation of ECM-receptor interaction pathways in PCOS, consistent with a fibrotic follicular environment.^34,37^

While our cell isolation workflow was not specifically designed to investigate theca and granulosa cells, other investigators have found dysregulated theca-immune cell signaling and distinct theca/granulosa transcriptomes in untreated PCOS, supporting the idea that normal immune activity is dysregulated at the ovarian level specifically. Overall, our results and those of others suggest a model wherein normal ovulation is a tightly controlled, acute inflammatory process driven by monocyte-derived cells and balanced by anti-inflammatory signaling. Stimulated follicles in PCOS patients, however, do not successfully activate the necessary inflammatory pathways, especially regarding monocyte and macrophage recruitment, and instead experience increased fibrosis and chronic inflammation.

While our study was limited by the use of exogenously FSH and LH-stimulated IVF patients, which may not capture all aspects of typical ovulation, our control data suggest that the fundamental inflammatory events of ovulation remain intact under IVF conditions.^28–32,39^ It is important to note that in our study, mature oocytes were retrieved from PCOS patients, so anovulation or failed oocyte-cumulus complex development is not responsible for the differences between PCOS and control follicles. Therefore our findings emphasize that the PCOS ovary does not simply fail to ovulate, but instead exhibits a fundamentally different immune state. Future studies may investigate the impact of specific androgen levels and/or resolution of the different PCOS phenotypes with regard to ovarian inflammation, as we matched patients with age and BMI but not androgen levels. Additionally, it may be worth investigating if tissue-targeted immune modulation could become a potential approach for improving ovulatory function or oocyte quality in PCOS.

In conclusion, our study presents a detailed immune cell atlas of the control and PCOS follicular environments, and suggest that chronic systemic inflammation in PCOS is accompanied by a failure to mount proper acute inflammatory pathways in the ovary. This immune dysregulation likely contributes to the ovulatory dysfunction in PCOS, and further investigation could inform targeted strategies to treat PCOS-associated infertility.

## METHODS

### Subject Recruitment and Inclusion/Exclusion Criteria

We selected 8 age and BMI matched control and PCOS patients from our consented and sampled patient pool UW-Madison IRB #2018-1247. The patients satisfied the Inclusion and exclusion criteria for the study (Table 1). 8 pairs of samples – TVOR appointment PBMC and Follicular Cells from each patient— were included in the study design. All patients had both peripheral blood and follicular fluid collected at the time of their transvaginal oocyte retrieval, following either an antagonist or Lupron overlap IVF protocol. Patient demographics and matching are in tables 1 and S1.

### Sample preparation

#### Mononuclear cell isolation

Peripheral blood mononuclear cells (PBMC) and follicular fluid mononuclear cells (FFMC) were separated from plasma and follicular fluid respectively by density gradient centrifugation and washed with RPMI + 10% fetal bovine serum. ACK lysis buffer was added to FFMCs to remove red blood cell contamination. Cells were counted, cryopreserved, and stored at -80 °C until experiments were conducted.

#### Enrichment for live cells

To ensure cell suspension viability after thawing, we enriched the samples for live cells by using a Dead Cell Removal Kit (Miltenyi Biotec, Bergisch Gladbach, Germany, cat# 130-090-101) with magnetic microbeads and Octomacs Magnetic separation columns (Miltenyi, cat # 130-042-109), following manufacturer’s protocol. Prior to our experiment, we tested this protocol on previously banked test samples and ensured that we produced high quality samples with > 90% viable cells. Experimental samples were further assessed using the Countess II (Invitrogen, Carlsbad, CA) upon submission to the UW Biotechnology Center Gene Expression Core and demonstrated 90-97% viability prior to library preparation and sequencing.

#### Multiplexing and Feature Barcode tagging of cells

Single cell suspension was prepared after checking for viability and count, starting with 1×10^6^ cells, suspended in Mg2+, Ca2+ and nuclease free buffer and incubated with Fc blocker (BioLegend, cat # 422301) for 10 mins at 40 ºC. The cell suspension was then incubated with appropriate multiplexing antibodies-TotalSeqB0252 antihuman Hashtag 1 (Biolegend cat #394631) for PBMC and TotalSeqB0252 antihuman Hashtag 2 (Biolegend cat #394633) for follicular cells for 30 mins at 40 C. The cell suspensions were washed and the tagged PBMC and follicular cell samples from each patient pooled into a single sample, to produce a total of 8 samples. The pooled samples were washed, resuspended in a labeling solution including TotalSeq B Feature Barcode (FB) antibodies (according to manufacturer’s instructions) and incubated for 30 mins at 4ºC. FB antibodies are listed in Table S2. After this final staining step, the cell suspension was washed, resuspended in a Mg2+, Ca2+ and nuclease free suspension buffer and submitted to the UW Biotechnology Center for library preparation and sequencing.

#### Library preparations and sequencing

Cell suspensions were submitted to UW Biotechnology Center where cell viability was further validated using the CountessTM II (Invitrogen). All samples met the initial quality control criteria and proceeded to single cell RNA library preparation and sequencing on the 10X Genomics platform. Libraries were prepared and downstream processing utilizing Illumina Novaseq X was performed by the University of Wisconsin Gene Expression Center in collaboration with the UWBC DNA Sequencing Facility, Madison, Wisconsin.

### CITEseq Analysis

#### Single Cell Transcriptomics Data Preprocessing

Sequence data were processed with CellRanger v6.1.2. scRNA sequences were aligned to the *Homo sapiens hg38* assembly. Filtered gene matrices were analyzed in R using Seurat v5.1.0 except where otherwise specified. Doublets were annotated using ScDblFinder (Bioconductor) and removed, and then all eight samples were merged into a single Seurat object. For initial quality control, cells were retained that had between 200 and 10000 features, lower than 15% mitochondrial transcripts, total RNA count below 150000, and were annotated as singlets by scDblFinder (Table S3). Counts were depth-normalized and log-transformed, the top 2000 variable features determined, and counts scaled. Principle component analysis was used for dimensionality reduction. A k-nearest-neighbor graph was constructed (FindNeighbors, FindClusters; Leiden clustering with resolution 0.8) and visualized using uniform manifold approximation and projection (RunUMAP).

#### Hashtag separation and tissue annotation

PBMCs were bound with HTO- 1 and FFMCs with HTO- 2. Hashtag expression levels were scaled and cells with HTO-1 expression > 6 and HTO-2 expression < 7 were annotated as PBMCs. Cells with HTO-1 expression < 6 were annotated as FFMCs, and cells with expression of both hashtags < 7 could not be reliably demultiplexed and were excluded. Demultiplexing was validated by the presence of the granulosa and theca clusters almost exclusively in the FFMC group.

#### Single-Cell Cluster Annotation

Clusters were annotated based on gene expression and canonical immune cell surface markers (read out by DNA-tagged antibodies) and using a combination of cell type marker genes as well as highly expressed genes identified by rank-sum tests (FindMarkers and FindAllMarkers). Genes used to determine cluster identities are found in the relevant figures. Cells were first clustered based on broad cell type and NK cells, follicular cells, and T cells were extracted as separate Seurat objects, subclustered, and annotated in finer detail before being merged back with the original object.

#### Differential gene expression and functional enrichment analysis

Differential gene expression analyses were carried out using Model-based Analysis of Single-cell Transcriptomics (MAST) implemented through Seurat FindMarkers and plotted with EnhancedVolcano. Heatmaps were created with ComplexHeatmap, with expression data pseudobulked via AggregateExpression. For KEGG pathway and Gene Ontology analyses, genes with log2FC > 0.25 and FDR > 0.05 (Benjamini-Hochberg) were analyzed using clusterProfiler’s enrichKEGG and enrichGO functions.

#### CellChatDB

Seurat objects containing control and PCOS follicular cells were converted to CellChat objects and processed using the CellChat v2 pipeline and CellChatDB.human database. Each object was processed separately and then combined for comparison. Individual signaling pathways were visualized using the chord layout of netVisual_Aggregate.

### Statistical analysis

All statistical analyses were performed in R, using Seurat for single-cell analysis or base R packages for other statistics. For differential gene expression, MAST was implemented through Seurat with additional Benjamini-Hochberg FDR correction for downstream analyses. For KEGG and GO analyses, the top 20 pathways/gene ontology terms by Bonferroni-corrected p-value are shown. For cell proportion comparisons, paired (for FFMCs and PBMCs between same patients) and unpaired (all other comparisons) two-tailed Student’s T-tests are used, with results considered significant at p < 0.05. Further details are provided in figure legends and table S4.

## Data Availability

Sequencing data and count matrices are deposited in the NCBI Gene Expression Omnibus/Sequence Read Archive (Bioproject ID PRJNA1299291). Differential gene expression analysis results for main-text comparisons as well as scripts for all analyses are available at https://github.com/staniclab/PCOS-CITE-Seq2025

## SUPPORTING INFORMATION

**Table S1. Individual patient demographics and fertility outcomes**

**Table S2. Antibodies used in CITEseq**

**Table S3. Quality control and cell counts by patient**

**Table S4. Statistical analysis of cell proportions by tissue and PCOS vs. control**

**Figure S1. Quality control metrics are consistent across patient samples**

**Figure S2. Sub-clustering of selected immune populations**

**Figure S3. Cell identity differences by tissue and PCOS status**

**Figure S4. Cell-specific differential gene expression by tissue**

**Figure S5. Inflammatory genes expressed by both PCOS and control follicular cells**

**Figure S6. Differential expression of PCOS and control follicles**

## ACKNOWLEDGMENTS

The authors thank other members of the Stanic Lab, as well as Jenna Schmidt and members of the Schmidt lab, for thoughtful comments. We also thank study coordinators and other staff at the UW Fertility Care clinic. AKW was supported by NICHD T32HD1013840. This study was also supported by a grant from the American Society for Reproductive Medicine to LGC and AKS as well as UW-Madison OBGYN development funds and NICHD K12HD000849-28 to AKS. This study was completed with assistance from the UW-Madison Biotechnology Center Gene Expression Center (Research Resource Identifier – RRID:SCR_017757) and the University of Wisconsin-Madison Biotechnology Center Bioinformatics Core Facility (Research Resource Identifier – RRID:SCR_017799).

## AUTHOR CONTRIBUTIONS

AKW analyzed the data and primarily composed the manuscript. SB prepared samples and conducted sequencing. EG and RH analyzed data. FLJ prepared samples. AKS and LGC supervised the project, acquired funding, and participated in manuscript editing.

