## Supplemental Tables 1-4; Supplemental Figures 1-6 for "Single-cell analysis of follicular fluid reveals dysregulation of ovulatory immune function in PCOS patients undergoing ovarian stimulation"

**SUPPORTING INFORMATION**

**Table S1. Individual patient demographics and fertility outcomes**

**Table S2. Antibodies used in CITEseq**

**Table S3. Quality control and cell counts by patient**

**Table S4. Statistical analysis of cell proportions by tissue and PCOS vs. control**

**Figure S1. Quality control metrics are consistent across patient samples**

**Figure S2. Sub-clustering of selected immune populations**

**Figure S3. Cell identity differences by tissue and PCOS status**

**Figure S4. Cell-specific differential gene expression by tissue**

**Figure S5. Inflammatory genes expressed by both PCOS and control follicular cells**

**Figure S6. Differential expression of PCOS and control follicles**

**Table S1. Individual patient demographics and fertility outcomes**

| **Sample/Patient** | **Age Range** | **BMI Range** | **Oocytes retrieved** | **Mature eggs** | **Transfer Attempted** | **hCG** | **Live Birth** |
| --- | --- | --- | --- | --- | --- | --- | --- |
| **Control A** | **26-30** | **20-24** | **17** | **14** | **No** | **n/a** | **n/a** |
| **PCOS A** | **26-30** | **20-24** | **8** | **3** | **No** | **n/a** | **n/a** |
| **Control B** | **36-40** | **25-29** | **20** | **20** | **Yes** | **+** | **Yes** |
| **PCOS B** | **31-35** | **25-29** | **28** | **18** | **Yes** | **+** | **Yes** |
| **Control C** | **31-35** | **30-34** | **15** | **13** | **Yes** | **+** | **Yes** |
| **PCOS C** | **31-35** | **30-34** | **22** | **20** | **Yes** | **+** | **No (Spontaneous Abortion)** |
| **Control D** | **31-35** | **35+** | **6** | **3** | **Yes** | **-** | **Not Pregnant** |
| **PCOS D** | **31-35** | **35+** | **9** | **8** | **Yes** | **+** | **Yes** |

**Table S2. Antibodies used in CITEseq**

|  | Clone | Type of conjugated antibody | Vendor |
| --- | --- | --- | --- |
| CD45 | HI30 | TotalSeq-B | BioLegend |
| CD45 RA | HI100 | TotalSeq-B | BioLegend |
| CD45 RO | UCHL1 | TotalSeq-B | BioLegend |
| CD3 | SK7 | TotalSeq-B | BioLegend |
| CD4 | RPA-T4 | TotalSeq-B | BioLegend |
| CD8 | SK1 | TotalSeq-B | BioLegend |
| CD14 | 63D3/M5E2 | TotalSeq-B | BioLegend |
| CD19 | HIB19 | TotalSeq-B | BioLegend |
| CD16 | 3G8 | TotalSeq-B | BioLegend |
| CD56 | 5.1H11 | TotalSeq-B | BioLegend |
| CD94 | DX22 | TotalSeq-B | BioLegend |
| CD103 | Ber-ACT8 | TotalSeq-B | BioLegend |
| CD127 | A019D5 | TotalSeq-B | BioLegend |
| CD117 | 104D2 | TotalSeq-B | BioLegend |
| CD11b | ICRF44 | TotalSeq-B | BioLegend |
| CD11c | S-HCL-3 | TotalSeq-B | BioLegend |
| CD141 | M80 | TotalSeq-B | BioLegend |
| CD161 | HP3G10 | TotalSeq-B | BioLegend |
| CD123 | 6H6 | TotalSeq-B | BioLegend |
| HLA DR | L243 | TotalSeq-B | BioLegend |
| CD25 | BC96 | TotalSeq-B | BioLegend |
| CD279 (PD-1) | EH12.2H7 | TotalSeq-B | BioLegend |
| CCR7 or CD197 | 6H6 | TotalSeq-B | BioLegend |
| CD69 | FN50 | TotalSeq-B | BioLegend |
| CD34 | 581 | TotalSeq-B | BioLegend |
| CD49a | HMα1 | TotalSeq-B | BioLegend |
| CD49b | P1E6-C5 | TotalSeq-B | BioLegend |
| CD335 (NKp46) | 29A1.4 | TotalSeq-B | BioLegend |
| CD209 | 9E9A8 | TotalSeq-B | BioLegend |
| CD40 | 5C3 | TotalSeq-B | BioLegend |
| CD86 | IT2.2 | TotalSeq-B | BioLegend |
| CD274(PD-L1) | 29E.2A3 | TotalSeq-B | BioLegend |
| CD44 | BJ18 | TotalSeq-B | BioLegend |
| CD29 | TS2/16 | TotalSeq-B | BioLegend |
| Hashtag 1 | LNH-94; 2M2 | TotalSeq-B | BioLegend |
| hashtag 2 | LNH-94; 2M2 | TotalSeq-B | BioLegend |

**Table S3. Quality control and cell counts by patient**

| **Patient** | **Original Cell Counts** | **FFMCs Post-Quality Control** | **PBMCs Post-Quality Control** | **% Cells passing QC** |
| --- | --- | --- | --- | --- |
| **Control A** | 6216 | 976 | 3944 | 79 |
| **PCOS A** | 26224 | 6819 | 2401 | 35 |
| **Control B** | 15543 | 2512 | 6581 | 25 |
| **PCOS B** | 36894 | 432 | 7524 | 22 |
| **Control C** | 29094 | 411 | 9052 | 33 |
| **PCOS C** | 25508 | 717 | 12617 | 52 |
| **Control D** | 30557 | 2398 | 6955 | 31 |
| **PCOS D** | 12162 | 1503 | 3759 | 43 |

**Table S4: Statistical analysis of cell proportions by tissue and PCOS vs. control**

| **Figure** | **Comparison** | **n (per group)** | **P value** | **T statistic** |
| --- | --- | --- | --- | --- |
| **2C** | Classical Monocytes, FFMC vs PBMC | 8 | 0.025 | -2.83 |
|  | Macrophages, FFMC vs PBMC | 8 | 0.015 | 3.22 |
|  | Th22, FFMC vs PBMC | 8 | 0.0061 | 3.88 |
|  | Tissue resident cDC2, FFMC vs PBMC | 8 | 0.0065 | 3.88 |
| **2D** | Tissue resident cDC2, Control vs PCOS | 4 | 0.026 | -3.19 |

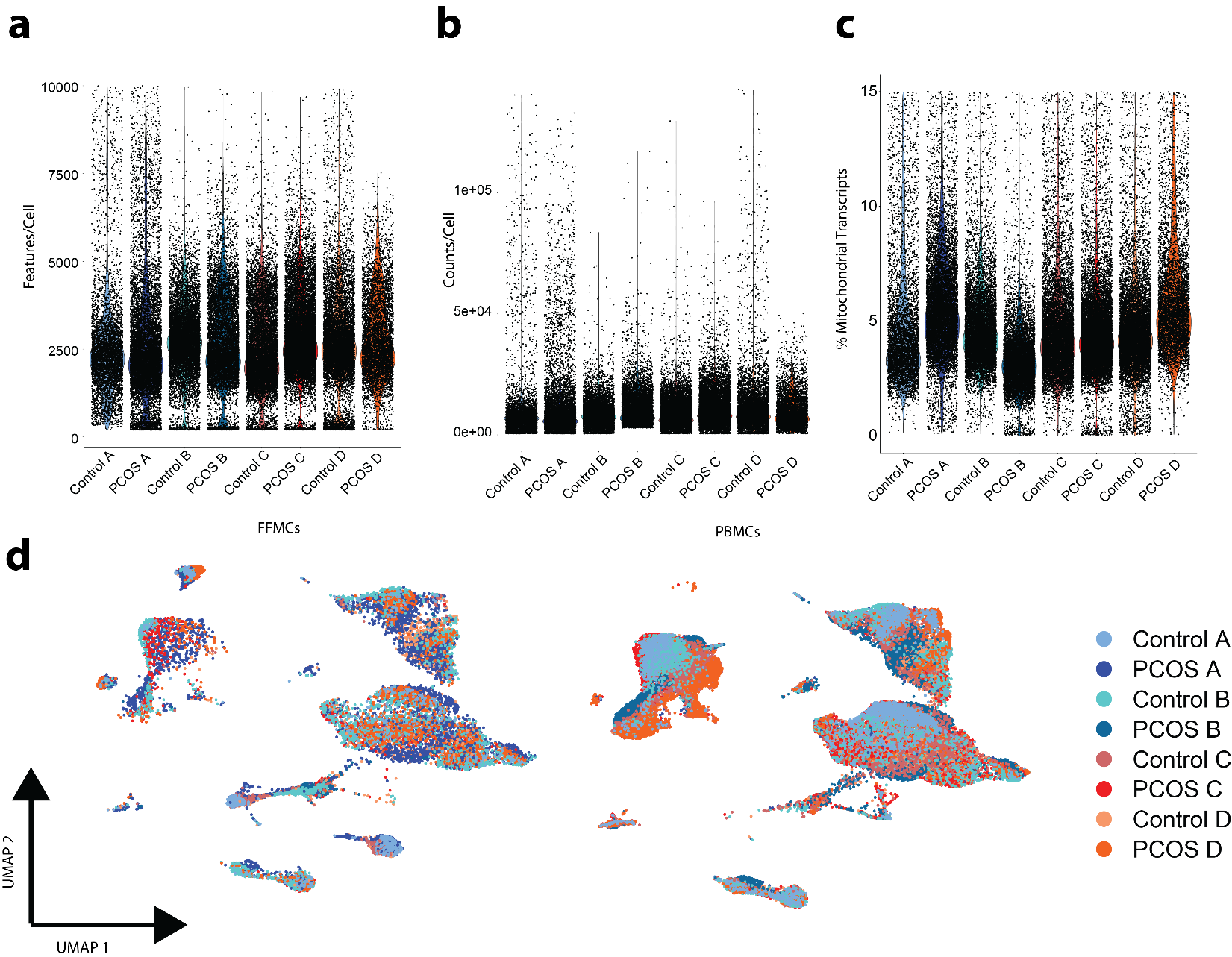
**Figure S1. Quality control metrics are consistent across patient samples.** A) Total features detected per cell for each patient following filtering. B) Total transcript counts detected per cell for each patient following filtering. C) Percent mitochondrial transcripts detected per cell for each patient following filtering. D) Distribution of follicular mononuclear cells and peripheral blood mononuclear cells by patient.

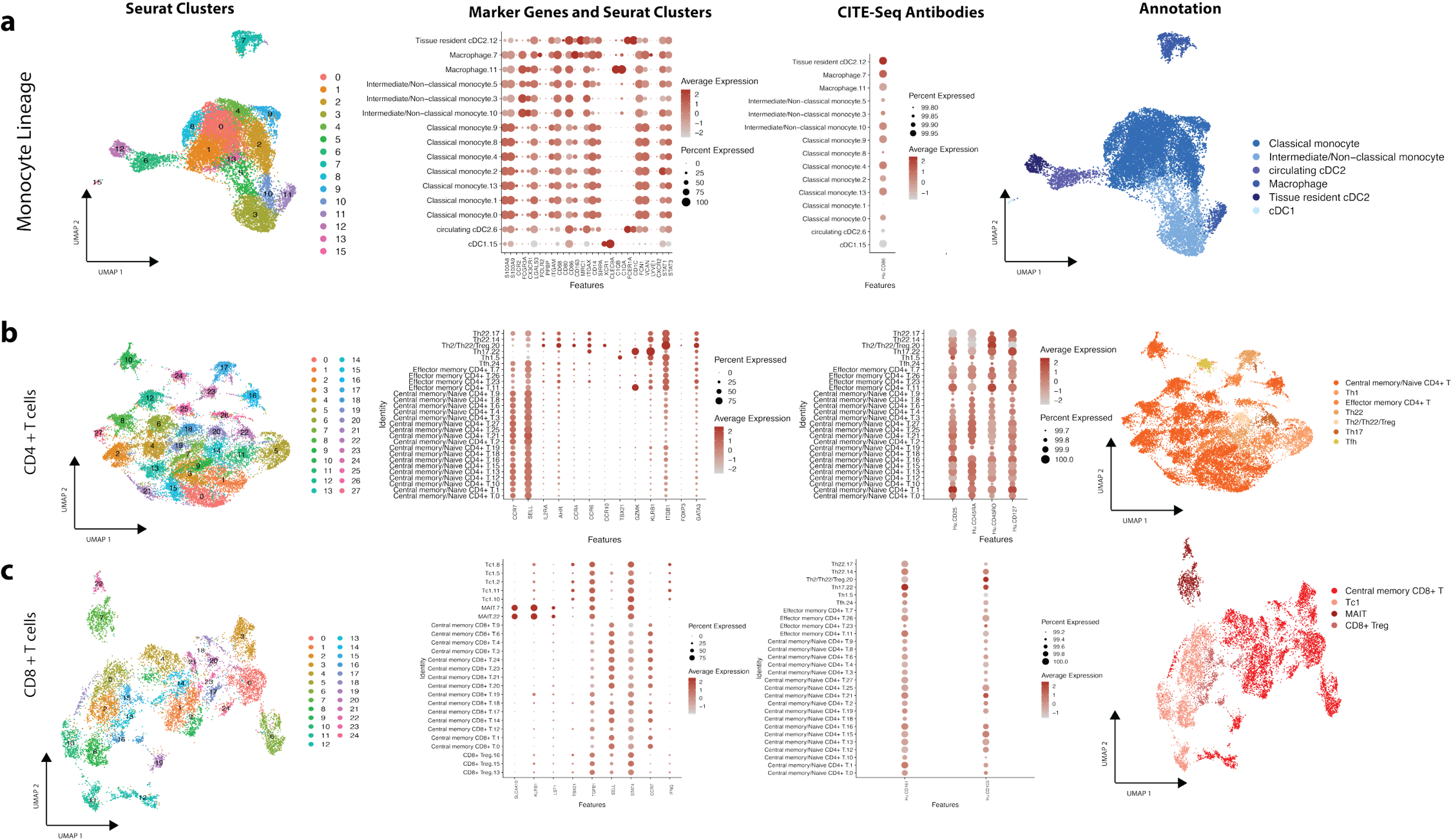

**Figure S2. Sub-clustering of selected immune populations.** Each of the selected cell subsets was individually clustered, annotated based on canonical immune marker genes and CITEseq antibody binding, and renamed before being reintegrated with the entire dataset.

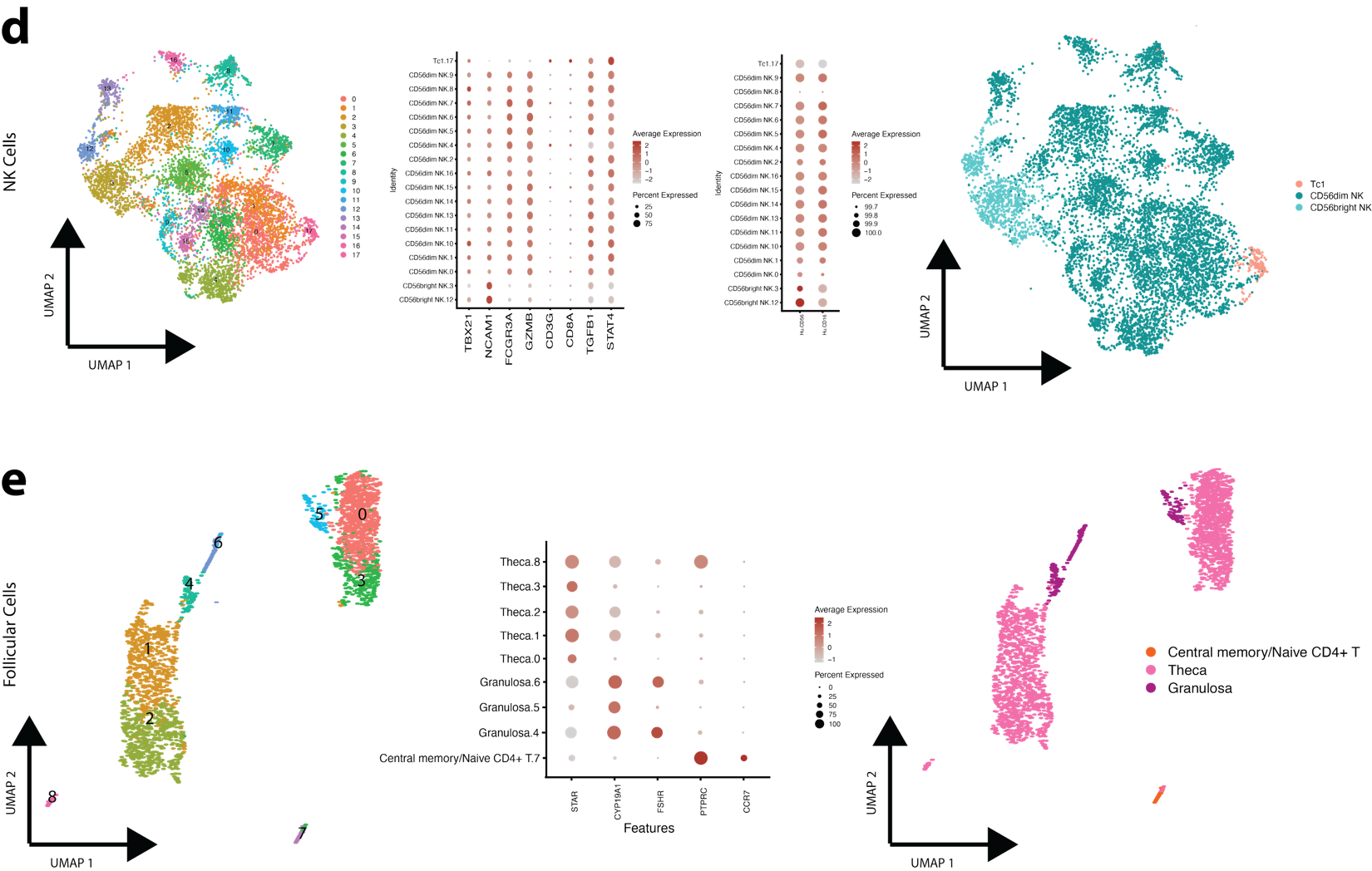

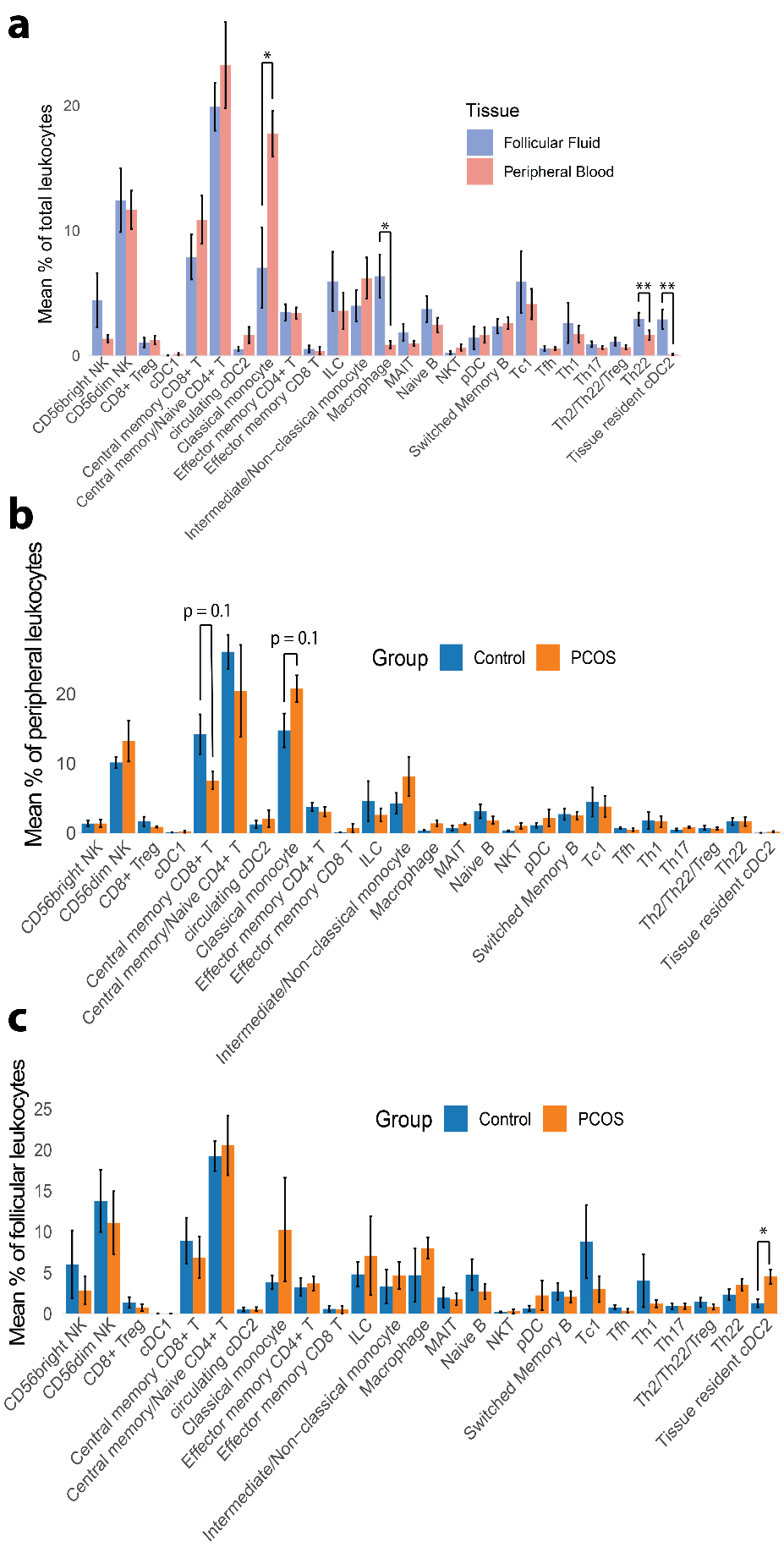
**Figure S3. Cell identity differences by tissue and PCOS status.** A) Mean % of total leukocytes (by patient) for each cell identity/annotation in follicular fluid vs peripheral blood. B) Mean % of all peripheral leukocytes (by patient) for each cell identity in PCOS versus control groups. C) Mean % of all follicular leukocytes (by patient) for each cell identity in PCOS versus control groups. Statistical analyses: Student’s t test, * p < 0.05, ** p < 0.01.

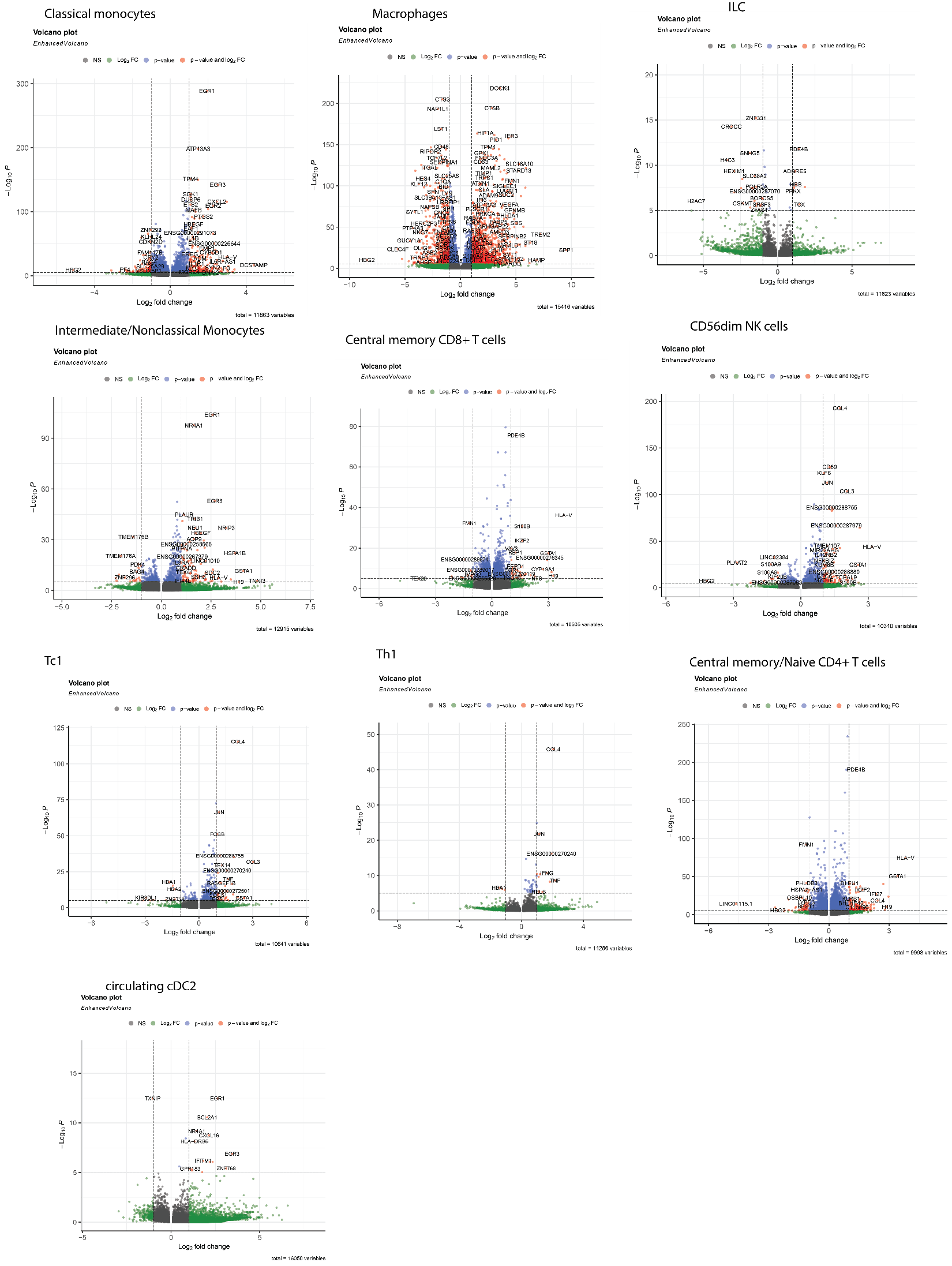

**Figure S4. Cell-specific differential gene expression by tissue.** Differential gene expression analyses of specific leukocyte cell identities, comparing follicular (positive log2FC) and peripheral (negative log2FC) cells.

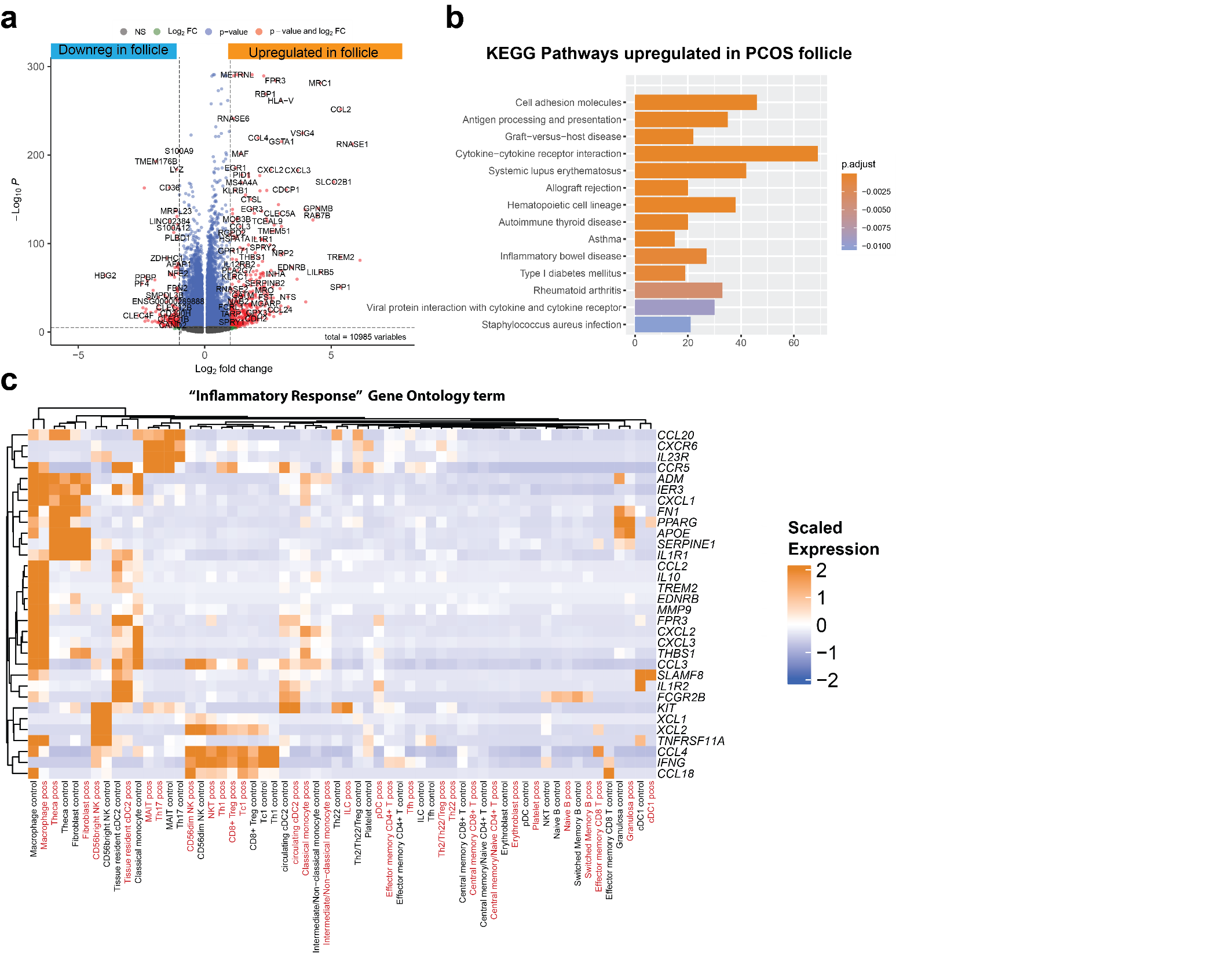
**Figure S5. PCOS follicles express an attenuated and broadly inflammatory transcriptional profile compared to both circulation and control follicles.** A) Differential gene expression analysis comparing genes upregulated in the PCOS ovarian follicle (positive log2FC) and in circulation in PCOS (negative log2FC). B) KEGG pathways most highly upregulated in PCOS follicles compared to circulation. C) Mean expression of genes found in the “inflammatory response” KEGG pathway that are upregulated by both PCOS and control follicular cells compared to their respective peripheral cells.

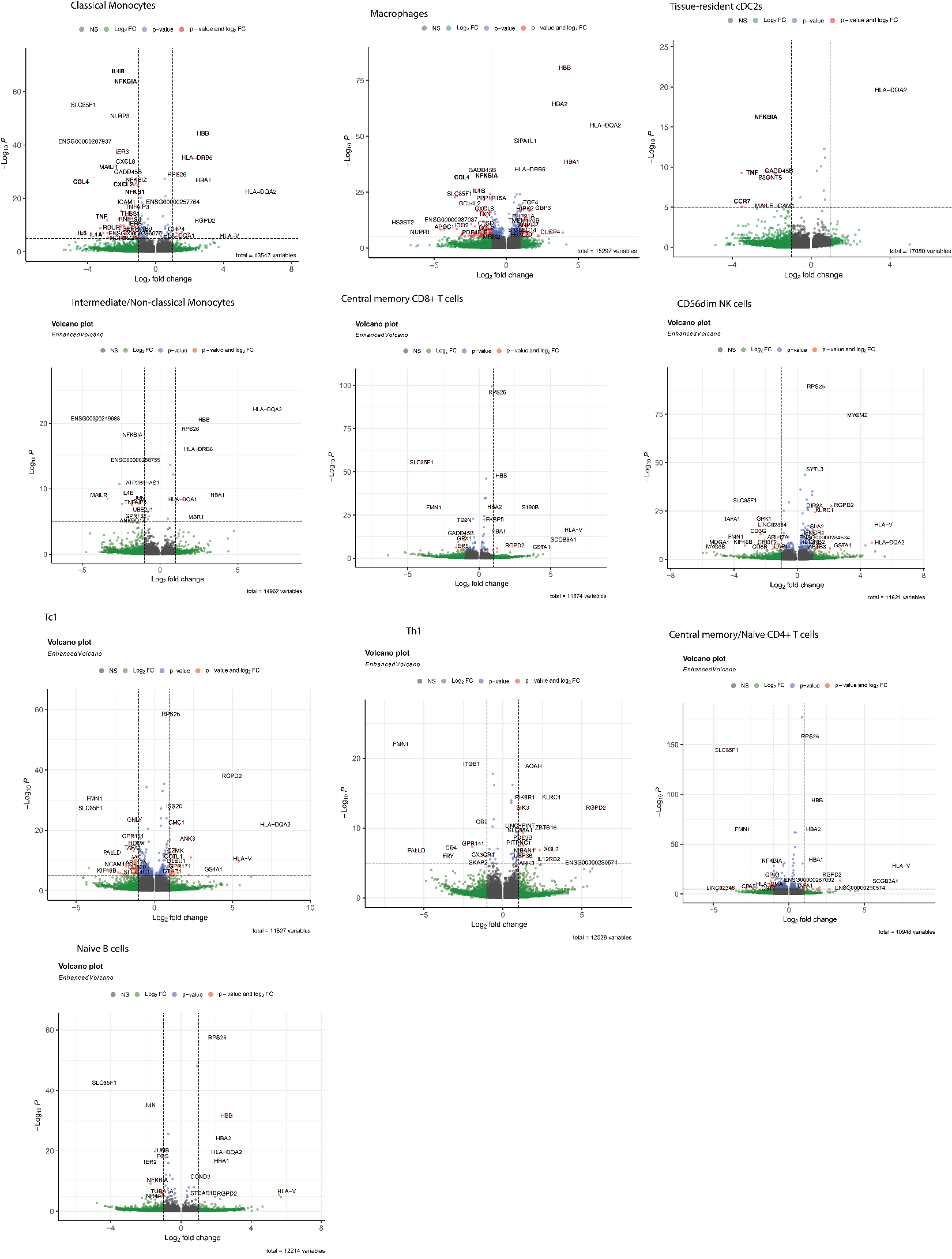

**Figure S6. Differential expression of PCOS and control follicles.** Differential gene expression analyses of specific leukocyte cell identities, comparing PCOS (positive log2FC) and control (negative log2FC) follicular cells.
